# A deep learning framework for comprehensive segmentation of deep grey nuclei

**DOI:** 10.64898/2025.12.16.25342423

**Authors:** Abhinabha Barat, Shridhar Singh, Ranjani Ramesh, Alberto Cacciola, Manojkumar Saranathan

## Abstract

**Background:** Deep grey matter structures such as the thalamus and basal nuclei are implicated in numerous neurological disorders, yet accurate segmentation of these structures from standard T1-weighted MRI remains challenging due to poor intra-subcortical contrast, long preprocessing pipelines, and fragmented toolsets.

**Methods:** We introduce THOMASINA a deep learning pipeline for comprehensive subcortical segmentation from standard T1-weighted (T1w) as well as white-matter-nulled (WMn) MRI. The method leverages labels derived from a recently published state-of-the-art multi-atlas segmentation method to train multiple 3D deep learning-based segmentation models including SwinUNETR, DiNTS, and SegResNet. All networks were trained on cropped volumes and tested on held-out and out-of-distribution datasets. For T1-weighted MRI, an additional synthesis step was used to generate WMn-like contrast prior to segmentation.

**Results:** SegResNet achieved the best performance (mean Dice = 0.89 on with in-domain test data, 0.85 on out-of-domain test data), outperforming DiNTS and SwinUNETR in both accuracy and robustness. It also had the highest mean, median, and minimum Dice and lowest SD in most nuclei compared to the DiNTS and SwinUNETR. Synthetic WMn contrast provided comparable segmentation to actual WMn images. The proposed networks reduced per-subject segmentation time to the order of seconds versus tens of minutes using traditional multi-atlas segmentation. THOMASINA also generalized well across field strengths, scanner vendors, and disease cohorts.

**Conclusions:** THOMASINA offers a fast, reproducible, and scalable solution for comprehensive subcortical segmentation from standard T1w MRI. By combining synthetic WMn contrast with state-of-the-art deep learning-based segmentation models, our method addresses key barriers to deployment and sets a foundation for biomarker discovery in clinical and population-scale imaging studies.

## Introduction

Deep grey matter structures such as the thalamus and its internal subdivisions, and basal nuclei serve as critical hubs for motor planning, cognitive control, affective processing, and sensorimotor integration. Accurate segmentation of these subcortical structures is crucial for elucidating the role of these important structures in many neurological and neuropsychiatric disorders including Alzheimer’s disease ^1,2^, Parkinson’s disease, frontotemporal dementia ^3^, multiple sclerosis ^4,5^, and schizophrenia ^6^. Quantification of atrophy and volumetric asymmetry at a substructural level may yield potential biomarkers in both research and clinical scenarios.

However, segmentation of deep grey nuclei from standard clinical T1-weighted (T1w) MRI remains challenging. These structures exhibit low contrast against surrounding tissues, especially in 3T Magnetization Prepared Rapid Acquisition Gradient Echo (MPRAGE) images. The small size and shape variability of structures like habenula, and lateral/medial geniculate nuclei, further complicates the segmentation. While there are many popularly used packages like Freesurfer ^7^, FSL ^8^, and VolBrain ^9^ for T1 MRI-based segmentation, they also have specific limitations with respect to subcortical structures. Most of these segmentation tools produce a subset of subcortical structures (typically accumbens, amygdala, caudate, pallidum, putamen, and thalamus) and as a “whole” i.e. ignoring subdivisions. FSL has been shown to be more accurate than Freesurfer for some nuclei ^10,11^ but lacks thalamic nuclei segmentation while FreeSurfer has recently added a module for thalamic nuclei segmentation ^12^, requiring users to “mix and match” software to optimize performance.

Thalamus-optimized multi-atlas segmentation (THOMAS) ^13^ leverages the improved intra-thalamic contrast from white-matter-nulling (WMn) to segment 12 thalamic nuclei. Unlike conventional MPRAGE where cerebrospinal fluid (CSF) is suppressed, WMn-MPRAGE nulls white-matter ^14^. THOMAS was recently adapted for conventional T1 i.e. CSF-nulled contrast using a simple preprocessing step called Histogram-based polynomial synthesis (HIPS) that synthesizes WMn-like images from T1-contrast MRI prior to THOMAS segmentation ^15^. Both THOMAS and HIPS-THOMAS are, however, restricted to thalamic nuclei. None of the popularly used methods, to date, segment structures like the claustrum, red nucleus or the internal/external subdivisions of the globus pallidus which is critical for DBS targeting, necessitating the use of specialized tools or manual segmentation. A unified software tool that comprehensively segments deep grey nuclei would bolster repeatability and reproducibility efforts as well as for minimize overheads in installation and maintenance.

Deep learning-based segmentation methods have revolutionized other areas of medical image processing and have been attempted for thalamic segmentation starting with the work of ^16^ who used cascaded multi-planar 2D U-nets for segmenting WMn-MPRAGE data by first segmenting the whole thalamus with one network and then sub-dividing that using another network to obtain constituent nuclei. To adapt this to conventional MPRAGE contrast, ^17^ proposed a synthesis-segmentation cascade which first synthesized WMn-like images from T1-contrast MRI and then segmented them, all using 3D U-nets. This was also replicated in the DeepTHALAMUS implementation of Ruiz-Perez et al. ^18^. One of the problems with synthesis-based methods is suboptimal generalization to out-of-domain (OOD) datasets especially in the synthesis step. A recent comparison of methods ^19^ showed the synthesis-segmentation CNN approach to be suboptimal when applied to new scenarios like 7T MRI, presumably due to the increased B1 inhomogeneity, despite N4 bias correction. HIPS-THOMAS on the other hand was shown to handle these new scenarios robustly.

Recently, HIPS-THOMAS was extended to include more deep grey nuclei in a method called sTHOMAS ^20^ and shown to outperform Freesurfer and FSL-FIRST for segmentation of subcortical structures. In addition to 12 thalamic nuclei, sTHOMAS also segmented all basal nuclei including globus pallidus interna and externa, amygdala, red nucleus, and the claustrum. However, the process of extending HIPS-THOMAS to subcortical structures in sTHOMAS increased the processing times due to the larger crop size needed to cover these new structures (especially amygdala and claustrum). Processing times of 30-40 min can still be prohibitive when analyzing large databases like the UK Biobank with 10s of 1000s of subjects. Furthermore, the method’s reliance of nonlinear registration makes it susceptible to failure in brains with enlarged ventricles, often occurring in dementia and in older individuals.

In this work, we present THalamic and Other Medial-forebrain Areas Segmented with Innovative Neural-network Architectures (THOMASINA), a modular and unified deep learning framework for ultrafast, robust, and accurate segmentation of deep grey subcortical structures from conventional T1w MRI. Building upon high-quality sTHOMAS labels, THOMASINA integrates both HIPS-transformed T1 MRI and WMn-MPRAGE data to train multiple state-of-the-art segmentation networks. Our goals are trifold: (1) To train a deep learning pipeline for subcortical structure segmentation using sTHOMAS labels obtained from HIPS-transformed T1 MPRAGE and WMn-MPRAGE images across multiple vendors, resolutions, and field strengths (2) To benchmark multiple deep learning segmentation models for accuracy, robustness, and generalizability (3) To analyze contrast and scanner field strength effects on model performance (e.g. T1 vs WMn, 3T vs 7T)

## Materials and Methods

### 2.1 Datasets and Labeling

For training and testing all networks, a cohort of 317 brain MRI volumes drawn from multiple sites, scanners, and research datasets, including Amsterdam Ultra-high field Adult Lifespan Database (AHEAD, ^21^), Human Connectome Project Young Adult (HCP-YA), and GE/Siemens/Philips 3T imaging studies acquired inhouse on healthy subjects was used. Imaging parameters included 1 mm isotropic MPRAGE scans at 3T (inhouse), 0.7mm isotropic MPRAGE scans at 3T (HCP) and 7T (AHEAD). For 7T AHEAD data, the T_1_ maps were used to synthesize WMn contrast images using the formula 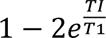 where TI was set to 650 ms for WM nulling. T1w MRI data from Alzheimer’s disease neuroimaging initiative (ADNI) database was used for out-of-domain testing (OOD, n=24) as well as a classification task with 380 cases divided into early mild cognitive impairment EMCI (n=110), late mild cognitive impairment LMCI (n=90), Alzheimer’s disease AD (n=72) and healthy controls (n=108) and these datasets were completely isolated from all model training/validation.

Segmentation labels for all cases were derived using the sTHOMAS pipeline, with additional HIPS preprocessing used for T1w MRI. To ensure a balanced and representative training set, we included both native WMn contrast and HIPS-synthesized T1 contrast images. The HIPS synthesis step generates WMn-like images from T1 prior to multi-atlas segmentation and has been shown to significantly improve the accuracy for standard T1w MRI segmentation ^15^ (see also preprocessing section below). sTHOMAS segmentation generates 10 thalamic nuclei, 5 basal nuclei (nucleus accumbens, caudate, putamen, globus pallidus externa and interna), habenula, mammillothalamic tract, claustrum, red nucleus, and amygdala, totaling 20 structures per hemisphere (Table 1). Note that in several cases (in addition to all AHEAD subjects), both T1 and WMn contrast images were available for the same subject. If a T1 contrast data set was picked for testing, WMn-contrast data from the same subject was also included in the testing to avoid test-train contamination even though these were independent acquisitions. All labels were in individual subject space and were quality-checked using visual inspection and used for training targets for our deep learning models. The dataset was split into 255 subjects for training and 62 for testing (standard 80-20 train/test split), with deliberate vendor/field strength stratification.

**Table 1.**
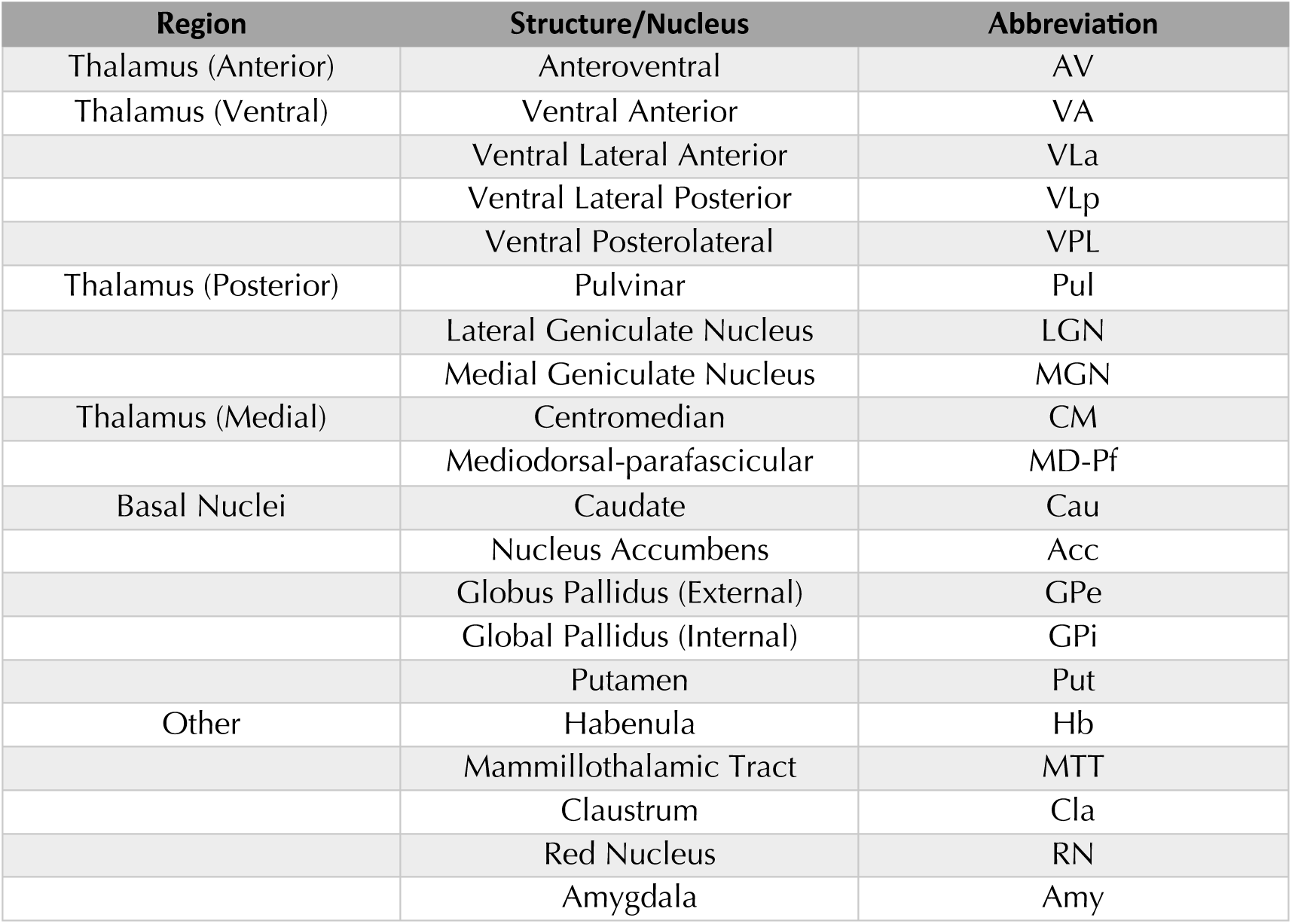
Segmentation labels comprise thalamic nuclei grouped as anterior, posterior, ventral and medial groups, basal nuclei (i.e caudate, putamen, globus pallidus interna and externa, nucleus accumbens), claustrum, red nucleus, amygdala and other deep grey matter nuclei (habenula, mammillothalamic tract). The notation matches the notation in sTHOMAS.

### 2.2 Preprocessing

The first step in sTHOMAS is automated cropping accomplished using affine registration using ANTs ^22^ to a template image and warping the crop mask from template space to input space. The cropped image is then N4-bias corrected to remove residual artifacts from B_1_ inhomogeneity. Cropping was introduced to reduce the size of the image and speed up the nonlinear registration step in THOMAS. In the case of deep learning, it additionally effects significant reduction in image size and memory requirements to enable use of 3D CNNs without memory or speed limitations.

For T1w MRI, an additional HIPS processing step was applied which uses a polynomial intensity mapping to synthetically convert T1 contrast MRI to WMn-like contrast. Such images are referred to as T1-HIPS in the manuscript as opposed to true WMn-contrast acquisitions. Finally, the images were reoriented to standard RAS orientation using *fslreorient2std* command of FSL.

### 2.3 Deep Learning Models

The overall workflow is illustrated in Figure 1. Examples of HIPS-synthesized T1 and native WMn images are shown to illustrate the contrast improvement of standard T1. To identify the most robust backbone for subcortical segmentation, we benchmarked three algorithms from the MONAI Auto3dSeg framework ^23^ under a unified preprocessing, training, and evaluation protocol:

- Swin-UNETR ^24^ – Transformer-based encoder–decoder combining Swin-Transformer blocks with U-shaped network architecture.
- DiNTS ^25^ – Differentiable Network Topology Search, a densely connected lattice based network.
- SegResNet ^26^ – Segmentation Residual Network, a U-net based CNN with 3D residual blocks.

**Figure 1.**
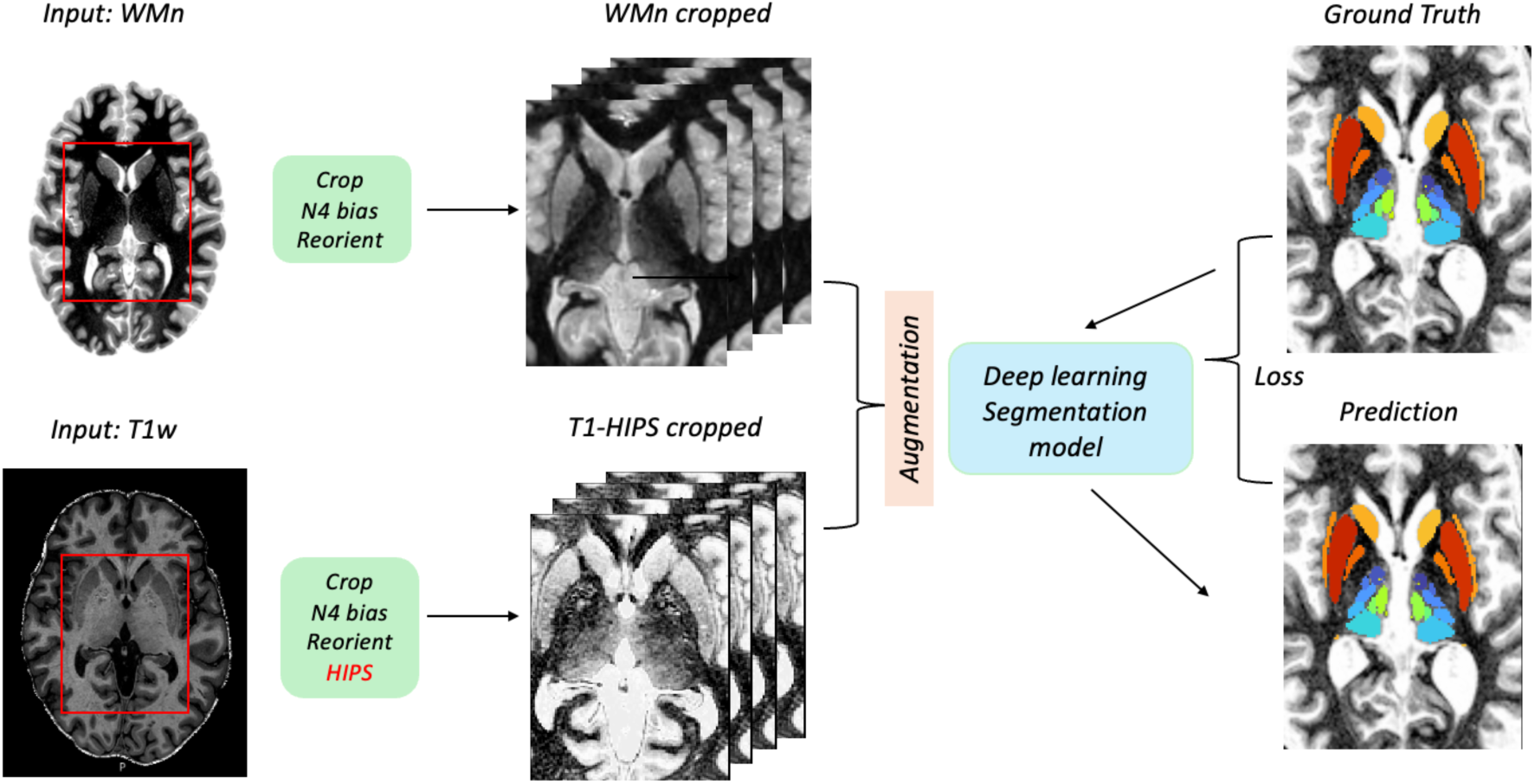
THOMASINA segmentation pipeline. T1-weighted MRI volumes are preprocessed using cropping to the region of interest. The cropped T1w volume is then passed through a Histogram-based polynomial synthesis (HIPS) step to synthesize a WMn-like image (T1-HIPS). Native WMn volumes undergo the same cropping regimen as the T1w MRI volumes (top left). Multiple segmentation models-Swin-UNETR, DiNTS, and SegResNet, are independently trained on the combined T1-HIPS and WMn contrast images. The resulting segmentations label 20 deep grey nuclei per hemisphere (bottom right).

All models were trained on the combined T1-HIPS and WMn datasets using a Dice + Cross Entropy loss, an AdamW optimizer, and dynamic patch sampling to balance foreground and background coverage. Data augmentation followed each network’s standard 3D medical imaging recipes and included random elastic deformation, Gaussian noise, intensity transforms, random scaling/zoom, flips, and cropping.

### 2.4 MONAI and Auto3dSeg workflow

All three networks-Swin-UNETR, DiNTS, and SegResNet-were trained using MONAI Auto3dSeg (v1.3). Auto3dSeg is MONAI’s self-configuring training framework that automates key steps of model development:

- **Data analysis and preprocessing**: It standardizes spacing, orientation, and intensity normalization, and automatically partitions the dataset into training/validation folds.
- **Architecture and hyper-parameter configuration**: The pipeline populates a set of candidate models (e.g., Swin-UNETR, SegResNet, DiNTS) with recommended patch sizes, learning rates, and optimizer schedules derived from dataset profiling.
- **Training orchestration**: Each candidate model is trained under their default augmentation recipes (e.g. random flips, rotations, elastic deformations, gamma/intensity shifts) and loss functions (Dice + Cross Entropy).
- **Ensembling and evaluation**: The best-performing checkpoints are optionally ensembled or exported for inference.

This approach ensures reproducible and hardware-efficient training across models while maintaining consistent data handling across architectures. Training time per model was approximately 3.5–4 hours on a single NVIDIA A100 GPU, using automatic mixed precision for efficiency. All test-time augmentations were disabled to preserve evaluation consistency, and inference was performed on an NVIDIA A100 GPU using automatic mixed precision for optimized speed and memory efficiency. The Auto3DSeg workflow used to deploy the SegResNet, DiNTS, and SwinUNET modules of THOMASINA requires at least a single GPU with more than 16GB RAM for both training and inference.

#### Initialization / pretraining

- SwinUNETR was initialized from public self-supervised weights distributed with MONAI Auto3DSeg. These weights originate from Swin-UNETR self-supervised pretraining on a large unlabeled CT corpus (learned with masked/inpainting, rotation, and contrastive proxy tasks) and are commonly used as a generic initialization prior to fine-tuning on target data (here, MRI)^27^
- DiNTS and SegResNet were trained from scratch with random initialization.

### 2.5 Evaluation Metric and Benchmarking Protocol

Model performance was evaluated on both the in-domain held-out test set and OOD ADNI cohort using the Dice Similarity Coefficient (DSC) as the primary segmentation accuracy metric. Special attention was given to evaluating performance on smaller and more challenging nuclei such as the centromedian (CM), habenula (Hb), and medial/lateral geniculate nuclei, as well as regions bordering the lateral ventricles, like the caudate nucleus, which are particularly prone to registration errors in traditional pipelines. We also conducted further analyses on the best performing model to assess robustness across image contrast types, comparing synthetic T1-HIPS and true WMn images, and across scanner field strengths (3T vs. 7T). Lastly, we also evaluated the performance of the best performing deep learning model of THOMASINA against sTHOMAS in Alzheimer’s disease stage discrimination on the ADNI cohort (i.e. EMCI, LMCI, and AD vs. HC). We used *pycaret* to perform the classification task, generate receiver operating characteristic (ROC) curves, and calculate area-under-the-curve (AUC) values based on the best performing classifier among 16 models including logistic and ridge regression, random forest classifier, linear discriminant analysis, and naïve Bayes.

## Results

### 3.1 Model Benchmarking and Architecture Selection

Figure 2 shows a heatmap of mean Dice scores for each of the deep learning models on the in-domain test set. Here, all 3D models performed well with SegResNet performing slightly better (mean Dice 0.89), compared to DiNTS (mean Dice 0.88) and Swin UNETR (mean Dice 0.86). Except for MTT and habenula, all structures achieve mean Dice of 0.8 or higher. Supplemental Table 1 lists the mean and SD of Dice, minimum Dice, and median Dice for all 3 models for each of the 20 nuclei for both hemispheres. In addition to the maximum mean and median Dice in most nuclei, SegResNet had overall lower SD and higher minimum Dice for most nuclei, indicating its stability and robustness compared to the other 2 models. Specifically, SegResNet had the highest mean and median Dice in 40/40 and 39/40 nuclei respectively, highest minimum Dice in 29/40 nuclei, and lowest variability (Dice SD) in 36/40 nuclei (Supplemental Figure 2).

**Figure 2.**
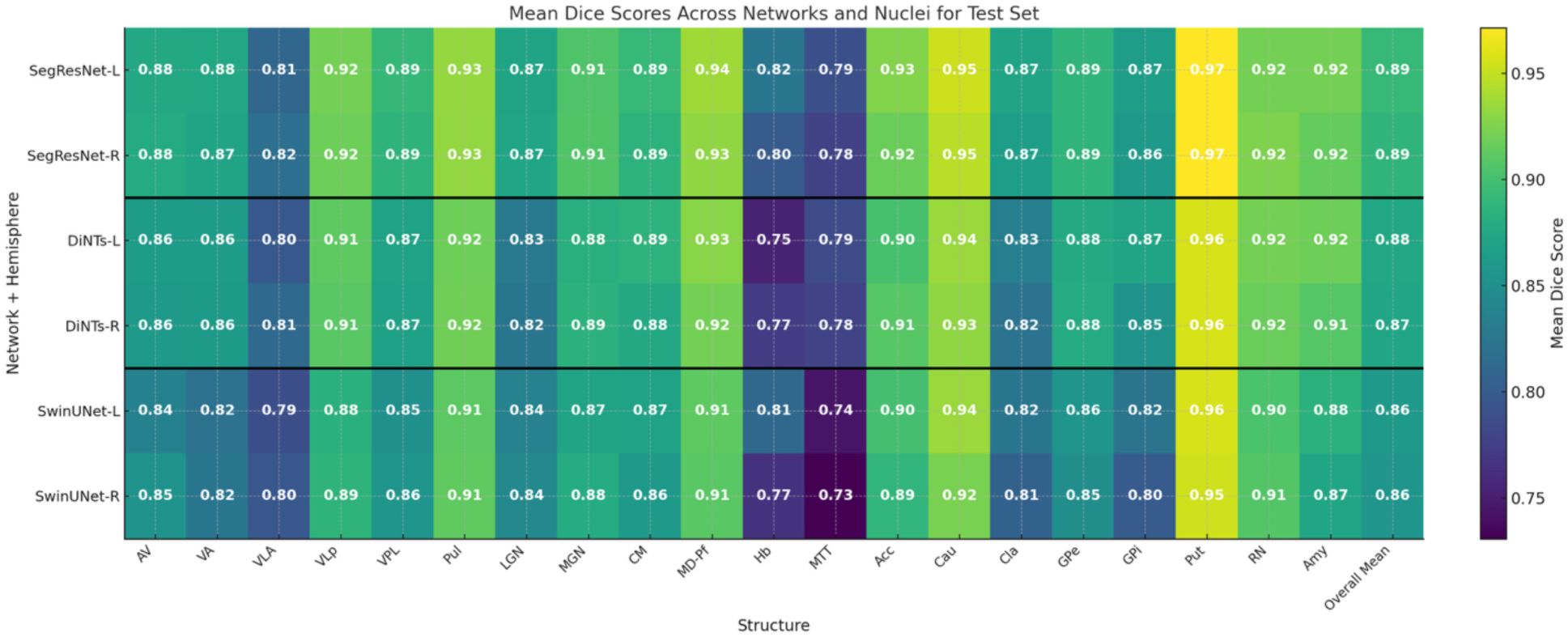
Heatmap of segmentation performance (mean Dice for each nucleus) for the three deep learning models using the in-domain test data (n=62) compared for left and right hemispheres. The mean across all nuclei is shown in the last column. SegResNet is slightly better than SwinUNet and DiNTs overall for all nuclei.

Figure 3 shows a heatmap of mean Dice scores for each of the deep learning models on the OOD ADNI test set. For this test set, a similar ranking was observed with SegResNet performing the best albeit with a slight reduction (∼0.04-0.05) in mean Dice across the board. Figure 4 shows boxplots of dice scores across all nuclei for SegResNet for the in-domain data set (top: left hemisphere, bottom: right hemisphere). Note that the order along x-axis is increasing nuclear volumes, showing (not surprisingly) that structures with larger volumes exhibit higher Dice with Putamen achieving the highest mean Dice of 0.94 and MTT, habenula, and ventral lateral anterior, the three smallest structures showing the lowest mean Dice of 0.78.

**Figure 3.**
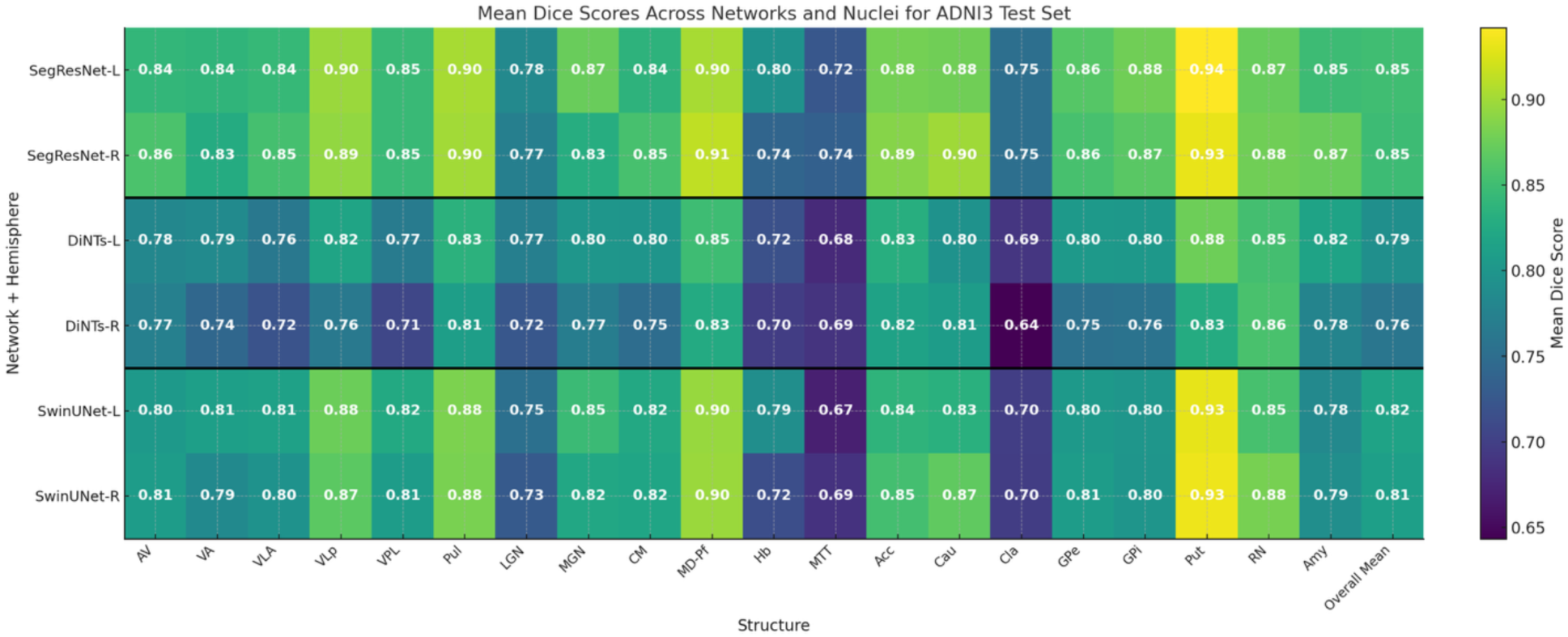
Heatmap of segmentation performance (mean Dice for each nucleus) for the three deep learning models using the out-of-domain ADNI test data (n=25) compared for left and right hemispheres. The mean across all nuclei is shown in the last column. SegResNet outperforms SwinUNet and DiNTs overall although showing a slight overall reduction compared to in-domain data (Figure 2). Unlike the in-domain data, DiNTS performed slightly worse than SwinUNET compared to SegResNet, indicating lesser ability to generalize.

**Figure 4.**
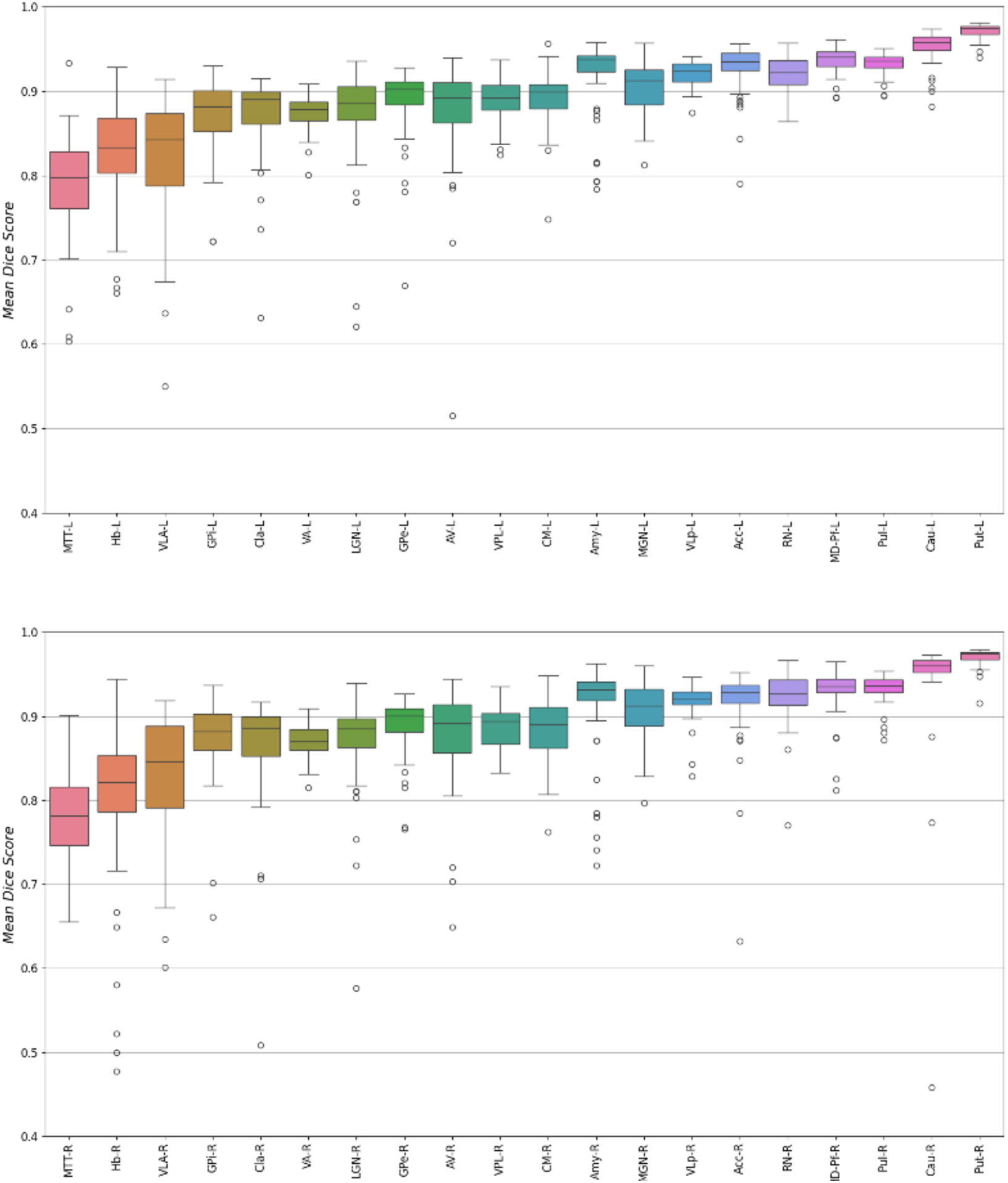
Boxplots of Dice scores for SegResNet for all 20 segmented deep grey nuclei for left (top) and right (bottom) hemispheres for the in-domain test data (n=62) with the structures ordered in the order of increasing size (left to right). SegResNet achieved high accuracy (∼0.9) across most structures, with minimal variance and limited outliers. Notably, even challenging structures such as the habenula (Hb) and mammillothalamic tract (MTT) achieved Dice of ∼0.8.

### 3.2 SegResNet Performance: Image contrast and field strength

Figure 5 stratifies SegResNet performance by image contrast (i.e. native WMn and T1-HIPS) and field strength (i.e. 3T vs 7T). There were no major differences observed indicating the ability of HIPS to successfully generate WMn-like contrast images comporting with the results of Vidal et al.^15^ as well as the results of Umapathy et al.^17^ which first demonstrated the superiority of WMn contrast over standard T1 contrast. Performance was also consistent across field strengths (mean Dice 0.86 @ 3T vs. mean Dice 0.87 @ 7T), attesting to SegResNet’s robustness.

**Figure 5.**
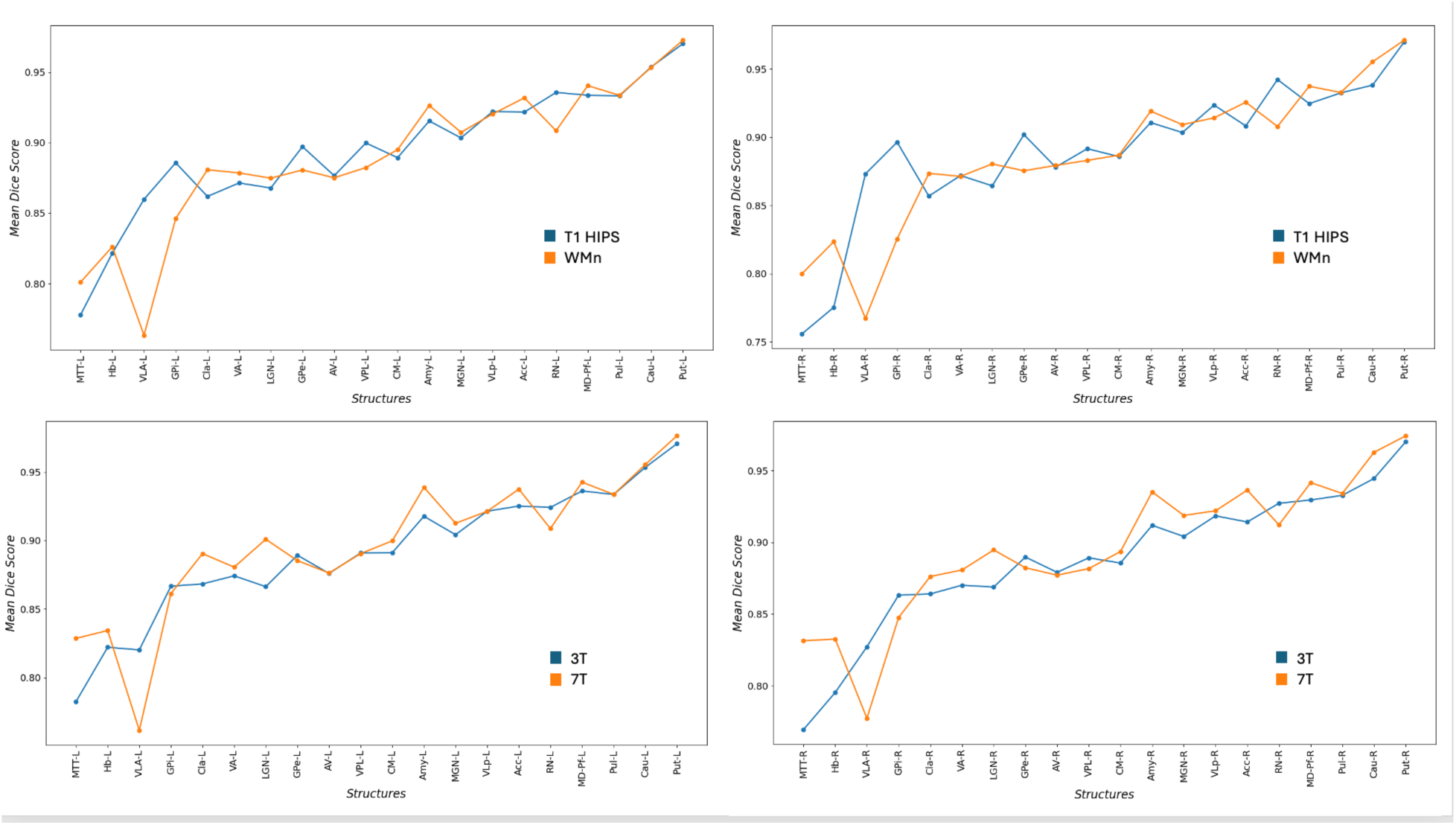
Mean SegResNet Dice stratified by image contrast i.e.T1 vs WMn (top row) and field strength i.e. 3T vs 7T (bottom row). The Dice values are highly concordant, indicating the robustness of the network across image contrasts and field strengths.

### 3.3 Performance in healthy control subjects and patients with enlarged ventricles

Figure 6 shows THOMASINA SegResNet (filled) and sTHOMAS (outline) segmentation outputs overlaid on a healthy control subject (top row) and a patient with frontotemporal dementia (FTD, bottom row). Notably, SegResNet maintained accurate anatomical delineation even in the presence of atrophy and modest ventricular enlargement in FTD, highlighting its robustness to structural variability. Figure 7 compares the performance of sTHOMAS (A,C) with THOMASINA (B,D) on two patients with FTD and severely enlarged ventricles. Note the failure of registration based sTHOMAS in the caudate nucleus (black arrows) whilst THOMASINA was able to segment the caudate nucleus successfully, indicating a potential benefit of this registration-free method even if accuracy is potentially comparable to sTHOMAS.

**Figure 6.**
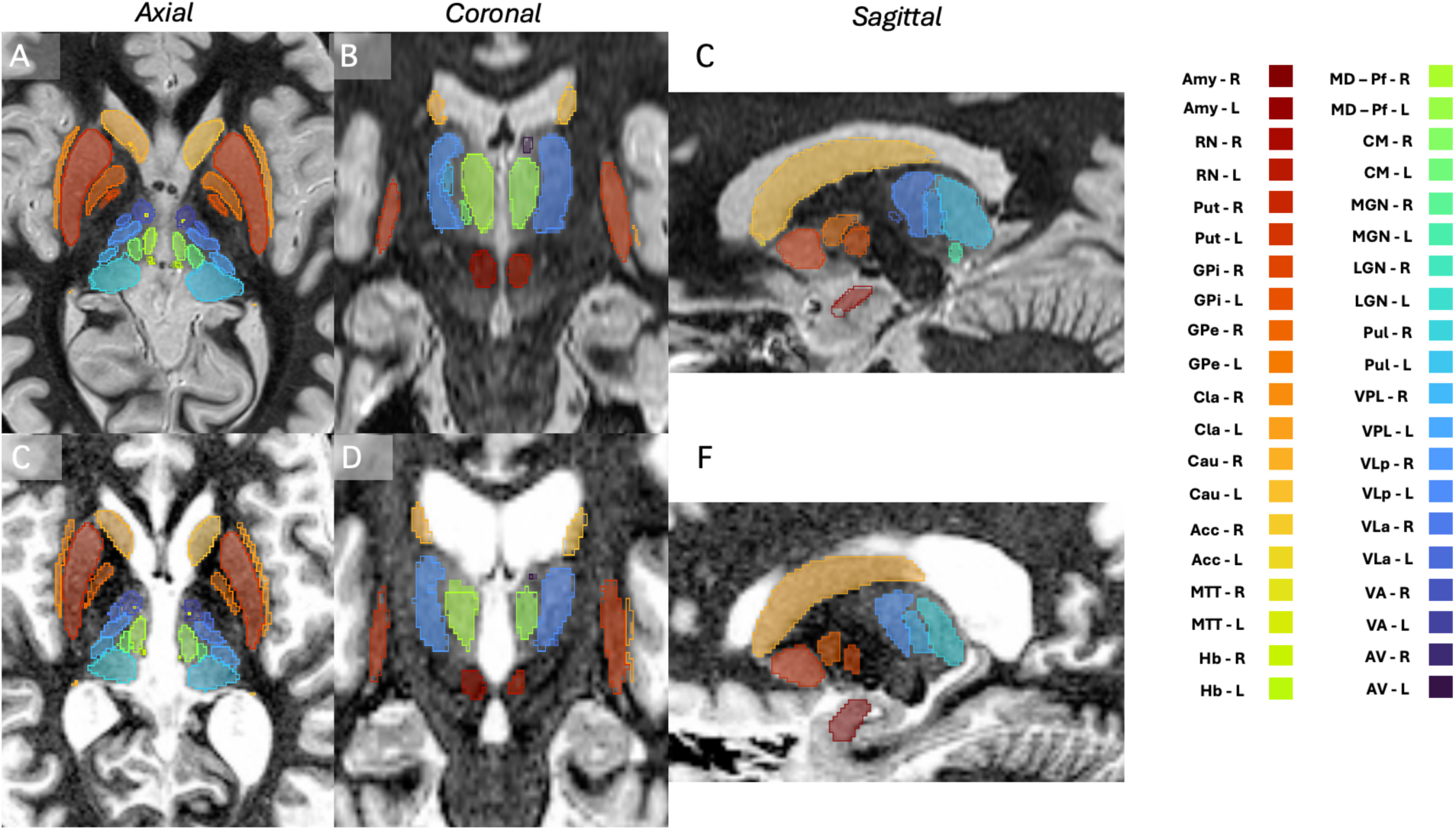
THOMASINA (solid) vs. sTHOMAS (outline) segmentation labels overlaid on T1-HIPS MRI data from a representative healthy control subject (top) and a frontotemporal dementia patient (bottom) shown in axial (A,C), coronal (B,D) and sagittal (C,F) planes. The color coding for the 20 labels generated per hemisphere are shown on the right.

**Figure 7.**
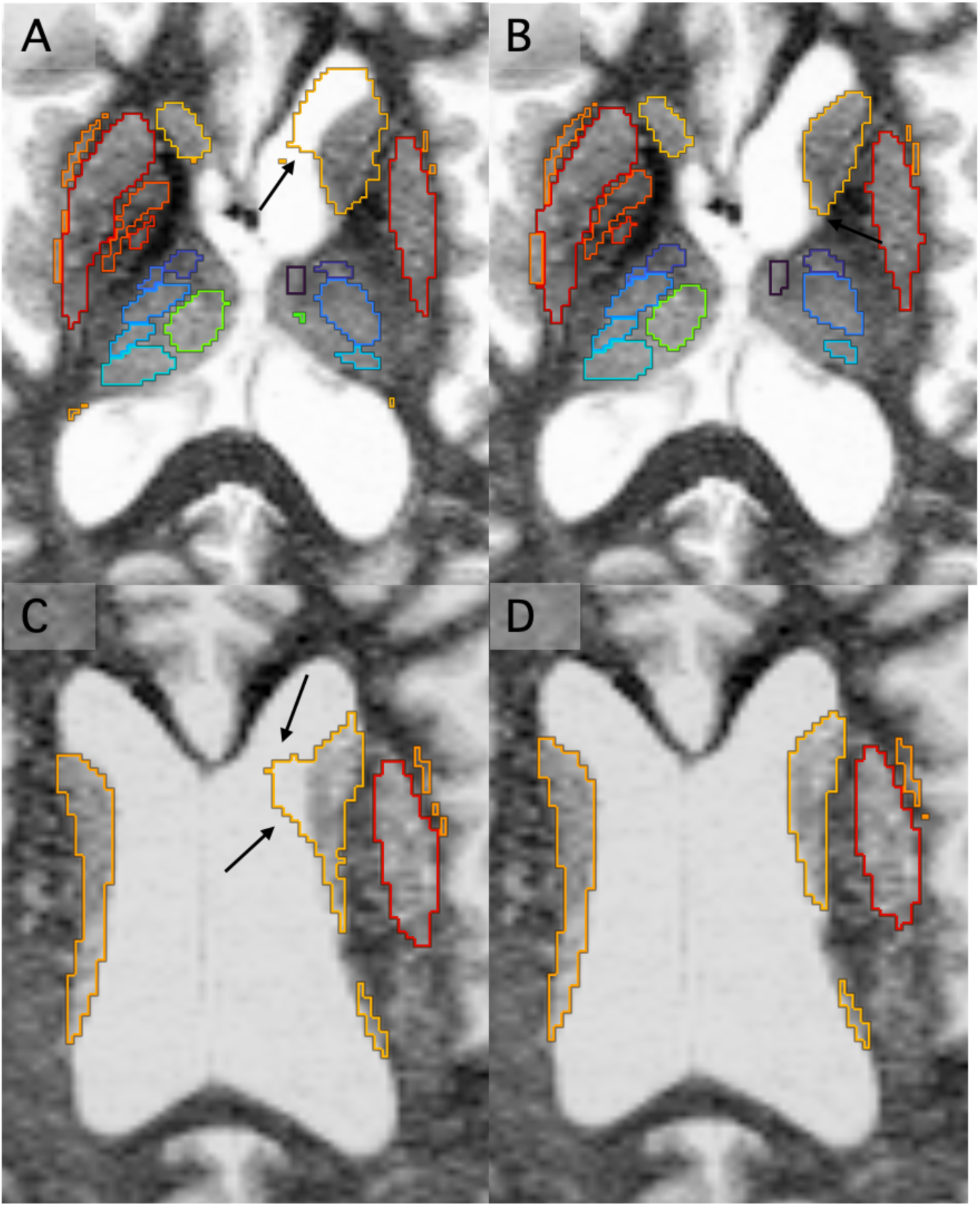
sTHOMAS (A, C) vs. THOMASINA (B, D) segmentations overlaid on T1-HIPS MRI data from two frontotemporal dementia patients with severely enlarged ventricles. Note the failure of sTHOMAS (black arrows in A, C) in the caudate region due to registration mismatches whilst THOMASINA does not display these failures.

### 3.4 ROC analysis

For all 3 classification scenarios (AD vs HC, LMCI vs. HC, and EMCI vs. HC), THOMASINA performed slightly better than sTHOMAS in most metrics notably AUC (0.90 vs 0.87 for AD, 0.78 vs 0.68 for LMCI, and 0.45 vs 0.60 for EMCI) indicating stability and robustness, especially for the critical EMCI where discrimination is difficult due to subtle atrophy. The ROC curves for all 3 scenarios are shown in Supplemental Figure 1.

## Discussion

We have developed THOMASINA, a robust, accurate, and efficient deep learning pipeline for segmenting subcortical structures from *both* standard T1-weighted MRI and WMn MRI. To our knowledge, this is the first deep learning-based method for comprehensive segmentation of deep grey nuclei including claustrum and subdivisions of the globus pallidus. By combining high-quality sTHOMAS labels, synthesizing WMn contrast from T1 MRI via HIPS, and using state-of-the-art deep learning architectures, THOMASINA achieves high accuracy and robustness whilst providing segmented outputs in under 10 seconds per scan. Our architecture benchmarking highlighted SegResNet as the most robust and accurate choice, performing well even in cases of enlarged ventricles such as patients with FTD, where registration-based THOMAS performs sub-optimally. THOMASINA also demonstrated robust performance across field strengths and image contrasts, attesting to its robustness.

WMn contrast significantly improves intrathalamic contrast as well as discrimination of the ventral boundaries against the abutting white matter tracks compared to standard T1w MRI contrast (from MPRAGE). This key insight in HIPS-THOMAS has been shown to improve segmentation accuracy even when using deep learning-based methods (e.g. ^17,18^). Previous attempts at deep learning-based methods for thalamic nuclei segmentation either focused either on WMn contrast which is very limited in scope^16^ or used a separate CNN to synthesize WMn-contrast from T1 MRI prior to segmentation^17,18^. This makes it vulnerable to failure due to variability in post processing algorithms of different vendors and higher B1 shading in 7T MRI. On the other hand, the HIPS-synthesis approach proposed in Vidal et al.^15^ and used by THOMAS/sTHOMAS hand offers a fast alternative and has been shown to be highly robust to scanner type or field strength. We had originally proposed this approach for thalamic nuclei segmentation (Barat et al. ^28^) which was later also used by Cabeza-Ruiz et al^29^ on a small data set. In our work, we expanded this idea to labels from sTHOMAS to enable a more comprehensive subcortical deep grey parcellation.

While accuracy of THOMASINA was comparable to sTHOMAS, the benefits of a deep learning-based method are two-fold-registration-free segmentation and speed. Enlarged ventricles, often seen in elderly healthy subjects as well as patients with AD, FTD and other forms of dementia result in suboptimal performance of sTHOMAS due to its critical dependence on nonlinear registration. By directly segmenting the input data, THOMASINA avoids this and demonstrated excellent performance even in cases with extremely large ventricles. Another key strength of THOMASINA is its ability to deliver comprehensive subcortical segmentation in under 10 seconds per subject on a single modern GPU. This represents an orders-of-magnitude improvement over widely used reference pipelines. For example, segmentation times of sTHOMAS, FreeSurfer and FSL-FIRST are typically 30-40 minutes, 4–8 hours, and 30–40 minutes respectively for subcortical segmentation on standard CPU hardware. Note that Freesurfer generates whole brain segmentation including cortical structures and needs that first step to perform thalamic segmentation. THOMASINA’s vastly reduced segmentation times can be significant when performing large scale analysis of databases like the UK Biobank with hundreds of thousands of subjects, planned for the near future.

Our architecture benchmarking step provided practical insights. Transformer based models such as SwinUNETR were moderately accurate but less robust to noise and resolution shifts, while architecture search models (DiNTS) offered strong in-distribution performance but lower generalization. In contrast, residual CNNs such as SegResNet consistently offered a compelling balance of speed, stability, and accuracy, underscoring their utility for both research and clinical applications. Future work will also include newer state-of-the-art models such as attention-enhanced U-nets ^30^ and Mamba-bot nets^31^.

Our method had some drawbacks. We used the cropping algorithm of THOMAS/sTHOMAS which is based on affine registration to an internal averaged template with a pre-defined crop mask that is then warped to native space and used for cropping. This increases the pre-processing time (∼1-2 min depending on spatial resolution). Future versions could incorporate a deep learning-based cropping network making the pre-processing times much faster. The HIPS preprocessing step tends to amplify noise due to the cubic and quadratic terms in the remapping process. This could be ameliorated using denoising of T1w MRI data prior to application of HIPS which requires more careful investigation. To evaluate the accuracy of THOMASINA, we used sTHOMAS as a ground truth for Dice computation. THOMAS and sTHOMAS were already evaluated against manual segmentation ground truth labels for thalamic and basal nuclei respectively using two different datasets. Another recent study found THOMAS to be superior to Freesurfer and its combined T1-DTI variant in a rigorous comparison involving two different atlases and area-under-the-curve accuracy in disease classification. Generating new manual segmentation ground truth labels for our test dataset was beyond the scope of this project and we settled for the suboptimal use of sTHOMAS as ground truth. Given the suboptimal performance of THOMAS in cases with large ventricles and the better performance of THOMASINA in disease classification, this implies that the true accuracy (Dice) of THOMASINA would be likely higher using manual segmentation ground truth. Lastly, SegResNet via the MONAI implementation requires a GPU even for inference. This could hamper the use of our method on older CPU-only systems.

In conclusion, THOMASINA provides a fully automated, end-to-end pipeline that generalizes across vendors, field strengths, and patient populations, and delivers accurate and comprehensive subcortical segmentation outputs in under 10s.

## Data Availability

All data produced in the present study are available upon reasonable request to the authors

## Acknowledgments

MS would like to acknowledge funding from the National Institute of Biomedical

Imaging and Bioengineering (R01 EB032674). We would like to thank Arit Banerjee for providing the sTHOMAS processed ADNI data which were used for classification.

Data were provided [in part] by the Human Connectome Project, WU-Minn Consortium (Principal Investigators: David Van Essen and Kamil Ugurbil; 1U54MH091657) funded by the 16 NIH Institutes and Centers that support the NIH Blueprint for Neuroscience Research; and by the McDonnell Center for Systems Neuroscience at Washington University.

ADNI: Data used in preparation of this article were obtained from the ADNI database (adni.loni.usc.edu). As such, the investigators within the ADNI contributed to the design and implementation of ADNI and/or provided data but did not participate in analysis or writing of this report. A complete listing of ADNI investigators can be found at: http://adni.loni.usc.edu/wpcontent/uploads/how_to_apply/ADNI_Acknowledgement_List.p df. Data collection and sharing for this project was funded by the Alzheimer’s Disease Neuroimaging Initiative (NIH Grant No. U01 AG024904) and DOD ADNI (Department of Defense Award No. W81XWH-12-2-0012). ADNI is funded by the National Institute on Aging, the National Institute of Biomedical Imaging and Bioengineering, and through generous contributions from the following: AbbVie, Alzheimer’s Association; Alzheimer’s Drug Discovery Foundation; Araclon Biotech; BioClinica, Inc.; Biogen; Bristol-Myers Squibb Company; CereSpir, Inc.; Cogstate; EISAI Inc.; Elan Pharmaceuticals Inc.; Eli Lilly and Company; EuroImmun; F. Hoffmann-La Roche Ltd. and its affiliated company Genentech, Inc.; Fujirebio; GE Healthcare; IXICO Ltd.; Janssen Alzheimer Immunotherapy Research & Development, LLC.; Johnson & Johnson Pharmaceutical Research & Development LLC.; Lumosity; Lundbeck; Merck & Co., Inc.; Meso Scale Diagnostics, LLC.; NeuroRxResearch; Neurotrack Technologies; Novartis Pharmaceuticals Corporation; Pfizer Inc.; Piramal Imaging; Servier; Takeda Pharmaceutical Company; and Transition Therapeutics. The Canadian Institutes of Health Research is providing funds to support ADNI clinical sites in Canada. Private sector contributions are facilitated by the Foundation for the NIH (http://www.fnih.org). The grantee organization is the Northern California Institute for Research and Education, and the study is coordinated by the Alzheimer’s Therapeutic Research Institute at the University of Southern California. ADNI data are disseminated by the Laboratory for Neuro Imaging at the University of Southern California.

## Supplemental Material

**Supplemental Figure 1.**
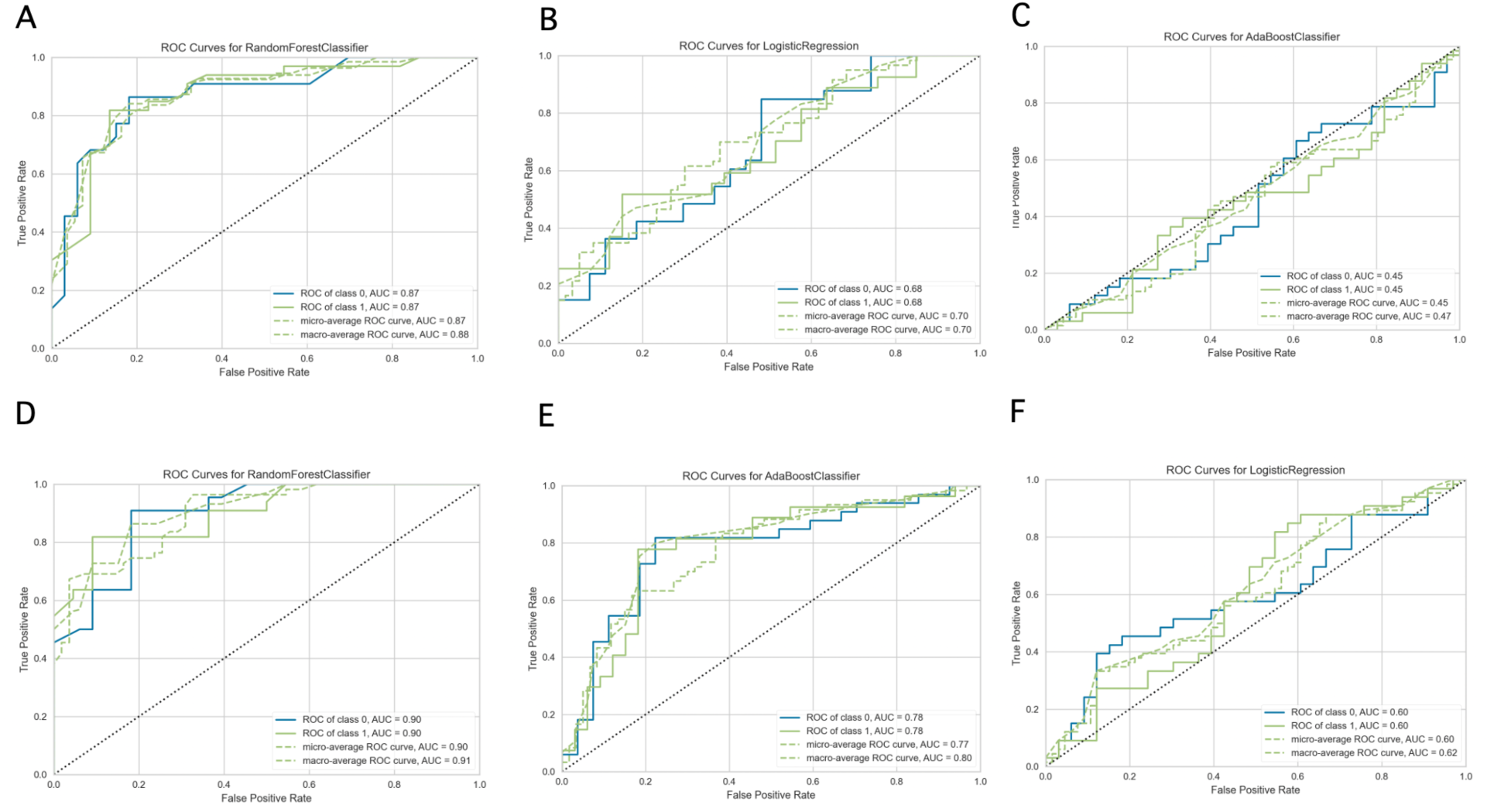
Receiver operating characteristic (ROC) analysis results for classification of Alzheimer’s disease (A, D), late mild cognitive impairment (B, E) and early mild cognitive impairment (C, F) vs. healthy controls for sTHOMAS (top row) vs. THOMASINA (bottom row). Area under the curve (AUC) values are significantly better for THOMASINA for both early (0.6 vs. 0.45) and late (0.78 vs. 0.68) mild cognitive impairment, and slightly better for Alzheimer’s disease (0.9 vs. 0.87) compared to sTHOMAS.

**Supplemental Table 1.** Mean, SD, Minimum and Median Dice coefficients for each nucleus for the three models.

|  | SegRes | SegRes | SegRes | SegRes | DiNTS | DiNTS | DiNTS | DiNTS | Swin Unet | Swin Unet | Swin Unet | Swin Unet |
| --- | --- | --- | --- | --- | --- | --- | --- | --- | --- | --- | --- | --- |
|  | mean | SD | min | median | mean | SD | min | median | mean | SD | min | median |
| L-AV | 0.87 | 0.06 | 0.51 | 0.89 | 0.86 | 0.09 | 0.37 | 0.88 | 0.84 | 0.09 | 0.39 | 0.86 |
| R-AV | 0.87 | 0.07 | 0.55 | 0.89 | 0.86 | 0.06 | 0.59 | 0.88 | 0.85 | 0.06 | 0.62 | 0.87 |
| L-VA | 0.87 | 0.02 | 0.80 | 0.88 | 0.86 | 0.02 | 0.80 | 0.87 | 0.82 | 0.04 | 0.62 | 0.83 |
| R-VA | 0.87 | 0.02 | 0.76 | 0.87 | 0.86 | 0.03 | 0.77 | 0.87 | 0.82 | 0.04 | 0.69 | 0.83 |
| L-VLA | 0.83 | 0.07 | 0.55 | 0.84 | 0.80 | 0.08 | 0.61 | 0.82 | 0.79 | 0.09 | 0.56 | 0.81 |
| R-VLA | 0.83 | 0.07 | 0.60 | 0.85 | 0.81 | 0.07 | 0.50 | 0.83 | 0.80 | 0.08 | 0.50 | 0.82 |
| L-VLP | 0.92 | 0.01 | 0.87 | 0.92 | 0.91 | 0.02 | 0.86 | 0.91 | 0.88 | 0.03 | 0.80 | 0.89 |
| R-VLP | 0.92 | 0.02 | 0.81 | 0.92 | 0.91 | 0.03 | 0.73 | 0.91 | 0.89 | 0.03 | 0.81 | 0.89 |
| L-VPL | 0.89 | 0.02 | 0.82 | 0.89 | 0.87 | 0.04 | 0.60 | 0.88 | 0.85 | 0.04 | 0.76 | 0.86 |
| R-VPL | 0.89 | 0.03 | 0.83 | 0.89 | 0.87 | 0.04 | 0.72 | 0.88 | 0.86 | 0.04 | 0.76 | 0.86 |
| L-Pul | 0.93 | 0.01 | 0.89 | 0.94 | 0.92 | 0.03 | 0.73 | 0.92 | 0.91 | 0.02 | 0.85 | 0.91 |
| R-Pul | 0.93 | 0.02 | 0.87 | 0.94 | 0.92 | 0.03 | 0.80 | 0.93 | 0.91 | 0.03 | 0.84 | 0.92 |
| L-LGN | 0.87 | 0.06 | 0.62 | 0.89 | 0.83 | 0.12 | 0.05 | 0.86 | 0.84 | 0.06 | 0.67 | 0.86 |
| R-LGN | 0.87 | 0.05 | 0.58 | 0.89 | 0.83 | 0.07 | 0.52 | 0.84 | 0.84 | 0.07 | 0.46 | 0.86 |
| L-MGN | 0.90 | 0.03 | 0.81 | 0.91 | 0.88 | 0.04 | 0.78 | 0.89 | 0.87 | 0.04 | 0.77 | 0.88 |
| R-MGN | 0.91 | 0.03 | 0.80 | 0.91 | 0.89 | 0.04 | 0.77 | 0.89 | 0.88 | 0.04 | 0.77 | 0.89 |
| L-CM | 0.89 | 0.03 | 0.75 | 0.90 | 0.89 | 0.02 | 0.80 | 0.89 | 0.87 | 0.03 | 0.77 | 0.87 |
| R-CM | 0.88 | 0.04 | 0.76 | 0.89 | 0.88 | 0.04 | 0.71 | 0.89 | 0.86 | 0.04 | 0.71 | 0.87 |
| L-MD | 0.94 | 0.02 | 0.87 | 0.94 | 0.93 | 0.02 | 0.81 | 0.94 | 0.91 | 0.02 | 0.83 | 0.92 |
| R-MD | 0.93 | 0.03 | 0.79 | 0.94 | 0.92 | 0.04 | 0.75 | 0.93 | 0.91 | 0.04 | 0.75 | 0.92 |
| L-Hb | 0.82 | 0.06 | 0.66 | 0.83 | 0.75 | 0.11 | 0.21 | 0.77 | 0.81 | 0.07 | 0.58 | 0.82 |
| R-Hb | 0.79 | 0.10 | 0.48 | 0.82 | 0.77 | 0.12 | 0.34 | 0.80 | 0.77 | 0.10 | 0.47 | 0.78 |
| L-MTT | 0.79 | 0.06 | 0.60 | 0.80 | 0.79 | 0.05 | 0.65 | 0.79 | 0.74 | 0.06 | 0.55 | 0.75 |
| R-MTT | 0.78 | 0.05 | 0.66 | 0.78 | 0.78 | 0.06 | 0.58 | 0.79 | 0.73 | 0.08 | 0.41 | 0.76 |
| L-Acc | 0.92 | 0.05 | 0.63 | 0.93 | 0.90 | 0.06 | 0.51 | 0.92 | 0.90 | 0.06 | 0.53 | 0.92 |
| R-Acc | 0.91 | 0.06 | 0.63 | 0.93 | 0.91 | 0.05 | 0.60 | 0.92 | 0.89 | 0.06 | 0.52 | 0.91 |
| L-Cau | 0.95 | 0.03 | 0.73 | 0.96 | 0.94 | 0.03 | 0.75 | 0.94 | 0.93 | 0.04 | 0.74 | 0.94 |
| R-Cau | 0.94 | 0.08 | 0.46 | 0.96 | 0.93 | 0.08 | 0.46 | 0.95 | 0.92 | 0.11 | 0.29 | 0.95 |
| L-Cla | 0.87 | 0.05 | 0.63 | 0.89 | 0.84 | 0.07 | 0.53 | 0.86 | 0.82 | 0.08 | 0.46 | 0.85 |
| R-Cla | 0.86 | 0.09 | 0.36 | 0.89 | 0.82 | 0.11 | 0.18 | 0.86 | 0.81 | 0.12 | 0.24 | 0.84 |
| L-Gpe | 0.89 | 0.04 | 0.67 | 0.90 | 0.88 | 0.04 | 0.75 | 0.89 | 0.86 | 0.04 | 0.70 | 0.87 |
| R-Gpe | 0.89 | 0.03 | 0.77 | 0.90 | 0.88 | 0.04 | 0.73 | 0.89 | 0.85 | 0.04 | 0.71 | 0.86 |
| L-Gpi | 0.87 | 0.07 | 0.39 | 0.88 | 0.86 | 0.07 | 0.44 | 0.88 | 0.82 | 0.07 | 0.43 | 0.84 |
| R-Gpi | 0.86 | 0.10 | 0.34 | 0.88 | 0.85 | 0.10 | 0.35 | 0.86 | 0.80 | 0.11 | 0.26 | 0.83 |
| L-Put | 0.97 | 0.01 | 0.93 | 0.97 | 0.96 | 0.02 | 0.84 | 0.96 | 0.96 | 0.02 | 0.90 | 0.96 |
| R-Put | 0.97 | 0.02 | 0.80 | 0.97 | 0.96 | 0.03 | 0.80 | 0.97 | 0.95 | 0.04 | 0.72 | 0.96 |
| L-RN | 0.92 | 0.02 | 0.86 | 0.92 | 0.92 | 0.03 | 0.85 | 0.92 | 0.90 | 0.03 | 0.85 | 0.91 |
| R-RN | 0.92 | 0.03 | 0.77 | 0.93 | 0.92 | 0.03 | 0.75 | 0.92 | 0.91 | 0.03 | 0.80 | 0.92 |
| L-Amy | 0.92 | 0.04 | 0.78 | 0.94 | 0.92 | 0.05 | 0.74 | 0.94 | 0.88 | 0.05 | 0.70 | 0.90 |
| R-Amy | 0.91 | 0.06 | 0.72 | 0.93 | 0.91 | 0.06 | 0.71 | 0.93 | 0.87 | 0.09 | 0.59 | 0.90 |
| Mean | 0.89 |  |  |  | 0.87 |  |  |  | 0.86 |  |  |  |

**Supplemental Figure 2.**
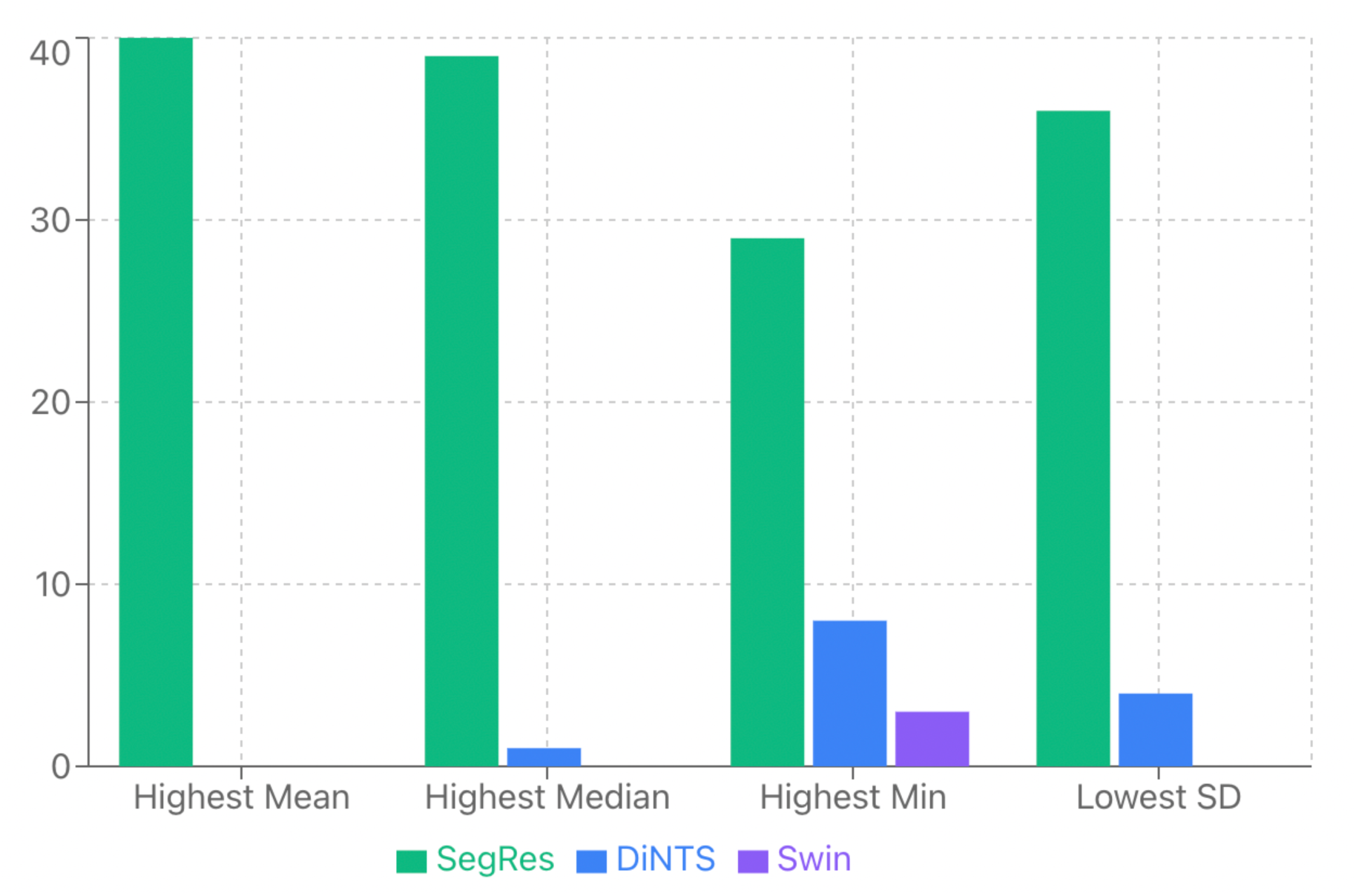
Comparison of performance between the 3 models: SegResNet had the highest mean, median, and minimum Dice and lowest SD in more nuclei than DiNTS and SwinUnet.

